# Prevention of Diabetic Foot Ulcer in High Risk Patient: The Role of Internist in Providing Adequate Foot Care

**DOI:** 10.1101/2021.02.07.21250589

**Authors:** Ida Ayu Kshanti, Nanang Soebijanto, Nadya Magfira, Marina Epriliawati, Jerry Nasarudin, Md Ikhsan Mokoagow

## Abstract

**Background and aim:** The awareness and practice of diabetes foot prevention by internist through routine examination and patient education remains less than optimal. This study aimed to evaluate the quality of care of internist in performing foot care in high-risk patients.

**Methods:** A cross sectional study was conducted in a tertiary care hospital in Jakarta, Indonesia. Type 2 diabetes mellitus subjects with high-risk foot complication were included in this study. Each subject filled-in a questionnaire investigating whether they had received information about proper foot care practice and whether they have ever had their feet examined by an internist at their present consultation. Multivariate logistic regression were performed.

**Results:** 368 patients were recruited, 130 of them (35.3%) treated by endocrinologists. 71.20% patients did not received any information on foot care and 54.08% patients did not received any foot examination. Foot care information was 1.6 times more frequently provided to patient with longer diabetes duration and 2 times more frequently provided to those who had history of diabetic foot ulcer. Meanwhile, foot examination was 1.5 times more frequently provided to those with longer diabetes duration and had a history of Lower Extremity Amputation. Compare to non-endocrinologist internist, endocrinologists tend to provide foot education and perform foot examination 2.2 to 2.5 times more frequently than non-endocrinologists.

**Conclusion:** Most of patients with high risk foot problems were not offered adequate foot care. It is necessary to develop strategies to improve the care and awareness among health professionals treating patients with diabetes especially internist.

## 1. Introduction

It is estimated that one in four people with diabetes will develop a diabetic foot ulcer (DFU) at least once in their life time.^1^ It is also common reason for hospital admission among people with diabetes and may lead to the amputation of the lower limb. A previous study showed that a lower limb is amputated due to diabetes every 30 s, and the average annual cost of diabetic foot is $8659 per patient.^2^ In Indonesia, it is estimated that there are 10.3 million people with DM, moreover 8.7% patients with DM had DFU with amputation rate 1.3%.^3-4^

It is well established in the literature that diabetes foot disease can be delayed or prevented through foot care.^5^ As recommended by International Working Group On The Diabetic Foot (IWGDF), key elements to prevent foot ulcers are; 1) identifying the at risk foot, 2) regularly inspecting and examining at risk foot, 3) foot care education, 4) ensuring appropriate footwear and 5) treating risk factor.^6^ People who were classified as high risk (history of ulceration, amputation, end-stage renal disease) were recommend to get foot screening and examination once every 1-3 months.^5^

Healthcare practitioners especially internists are actually aware of the consequences of diabetic foot.^7^ Unfortunately, the care of diabetic foot and its related condition are from optimal. Very few clinicians are treating the diabetic foot in a systematic, standardized method with proper risk categorization of foot complication.^8^ Auditing of all aspects of the service to ensure that local practice including quality of care of internist meets accepted international standards of care are necessary.^7^

In the context of improving the care of patients presenting with a diabetic foot, this study aimed to evaluate the quality of care of internist in performing foot care especially in high-risk patients.

## 2. Methods

### 2.1 Design and Settings

This is a cross sectional analytical study, which include 368 patients from Fatmawati General Hospital, Jakarta, Indonesia. Subjects were recruited from internal medicine department outpatient clinic between January-December 2016. At that time, in our clinic, there are 2 endocrinologists and 9 non-endocrinologist internists. All patients with type 2 diabetes mellitus will be assessed by endocrinologist once every 3 months, other than that time patient who came to our hospital for diabetes treatment can be seen by different non-endocrinologist internist according to internist on duty.

### 2.2 Sampling of patient

All patient with type 2 diabetes mellitus who had history of diabetic foot ulcer, and or history of low extremity amputation (LEA), and or history of chronic kidney disease (CKD); estimated glomerular filtration rate (eGFR) ≤ 60 mL/min were considered eligible for this study. Exclusion criteria for this study were patient with cognitive impairment, or in emergency situation or subjects with abbreviated mental state who cannot independently fill questionnaire. Patient were sampled using consecutive sampling. Ethics approval was obtained from the Ethical Committee for Medical Research, Fatmawati General Hospital.

### 2.3 Data collection procedures

Patient screening was done prior to the study to obtain the list of patients who were consider eligible for this study. These prospective subjects were then queuing in the polyclinic to underwent consultation to internist on duty. All of the internist during study time, were not know whether they patient will become study participants. After clinical assessment and consultation, a research team member will give oral and written information about the study. Patients who were willing to participate in this study were asked to sign a written informed consent.

Afterwards, the patients were asked to self-filled questionnaire investigating whether they had received information about foot care and whether they had had their feet examine by their prior internist during the latest consultation. Foot examination defined as physician perform anamnesis and or physical examination (seen, touch, etc.) to patient. All data concerning general medical history and laboratory data were collected by research member according to patient’s medical records and study forms specifically developed by the research committee. All data were transferred into a special clinical report form and were stored in the Integrated Center of Diabetes Care, Fatmawati General Hospital. The forms were checked for duplication before being entered into an electronic database.

### 2.4 Statistical analysis

Patient characteristic according to physician in charge, receiving foot education, performing foot examination were first compared using x^2^ test. To account for the multilevel nature of the data (patients clustered within physician or foot education or foot examination) and to control simultaneously for the possible cofounding effects of the different variables, we used multivariate logistic regression models. Results are expressed as prevalence ratio (PR) with their 95% confidence interval (95CI). In the analysis in correlates of foot education and examination, an OR> 1.0 indicates a higher likelihood of receiving the information or the examination. All the analysis was performed using STATA Version 12. This study had been approved by ethical committee of Fatmawati General Hospital 021/KPP/XI/2016

## 3. Results

### 3.1 Correlates of specialty of internist

Overall, 368 patients were recruited, of whom 130 (35.33%) treated by endocrinologist specialist. Among the latter, a previous history of foot ulcers was present in 270 patients (73.37%), 22 (5.98) had a previous history of LEA, and 158 patients (42.93) had CKD. The overall LEA event was 8.09% in patient with history of DFU. Patients characteristics according to internist specialty are reported in table 1. There is no correlation between internist specialty and patients’ characteristics.

**Table 1.**
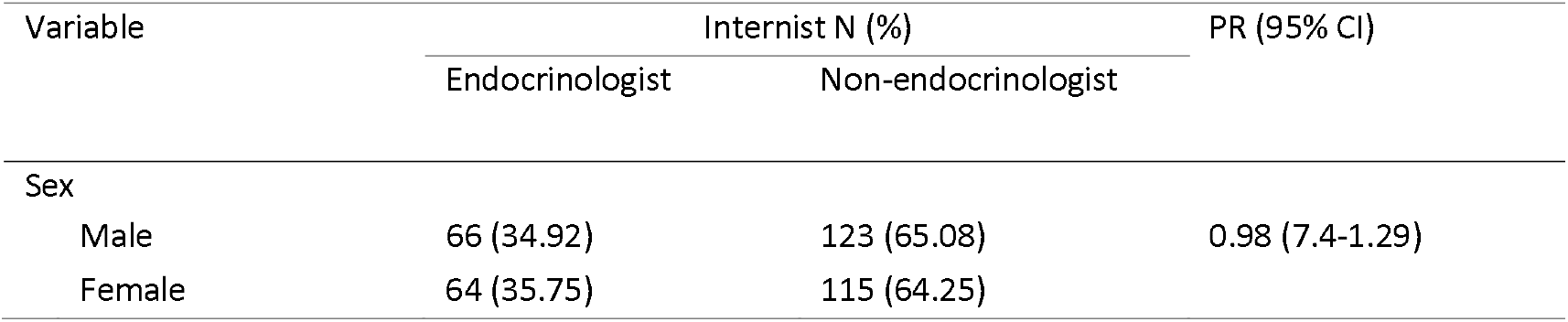
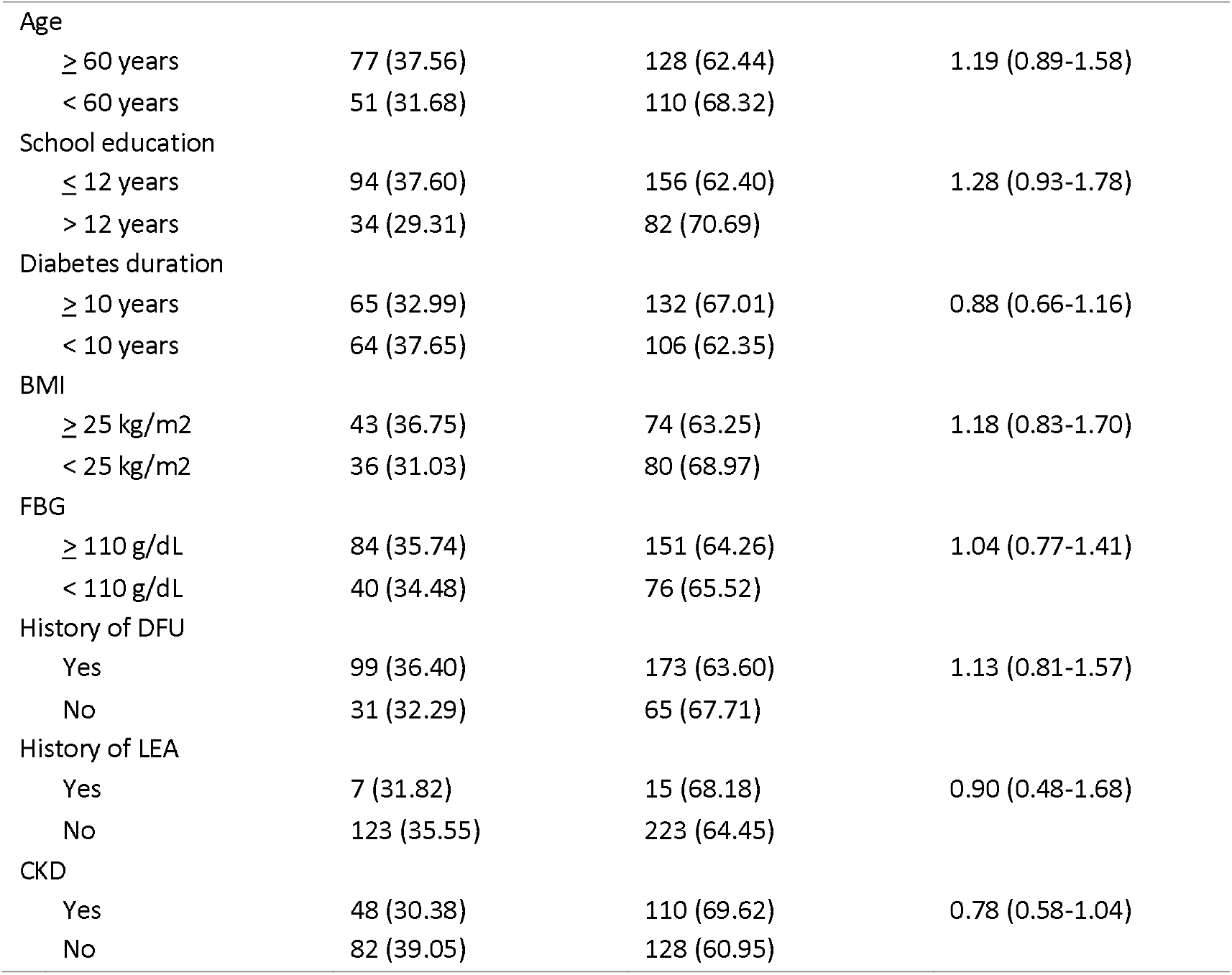
Patient Characteristics According To Internist Specialty

| Variable | Internist N (%) |  | PR (95% CI) |
| --- | --- | --- | --- |
|  | Endocrinologist | Non-endocrinologist |  |
| Sex |  |  |  |
| Male | 66 (34.92) | 123 (65.08) | 0.98 (7.4-1.29) |
| Female | 64 (35.75) | 115 (64.25) |  |
| Age |  |  |  |
| ≥ 60 years | 77 (37.56) | 128 (62.44) | 1.19 (0.89-1.58) |
| < 60 years | 51 (31.68) | 110 (68.32) |  |
| School education |  |  |  |
| ≤ 12 years | 94 (37.60) | 156 (62.40) | 1.28 (0.93-1.78) |
| > 12 years | 34 (29.31) | 82 (70.69) |  |
| Diabetes duration |  |  |  |
| ≥ 10 years | 65 (32.99) | 132 (67.01) | 0.88 (0.66-1.16) |
| < 10 years | 64 (37.65) | 106 (62.35) |  |
| BMI |  |  |  |
| ≥ 25 kg/m<sup>2</sup> | 43 (36.75) | 74 (63.25) | 1.18 (0.83-1.70) |
| < 25 kg/m<sup>2</sup> | 36 (31.03) | 80 (68.97) |  |
| FBG |  |  |  |
| ≥ 110 g/dL | 84 (35.74) | 151 (64.26) | 1.04 (0.77-1.41) |
| < 110 g/dL | 40 (34.48) | 76 (65.52) |  |
| History of DFU |  |  |  |
| Yes | 99 (36.40) | 173 (63.60) | 1.13 (0.81-1.57) |
| No | 31 (32.29) | 65 (67.71) |  |
| History of LEA |  |  |  |
| Yes | 7 (31.82) | 15 (68.18) | 0.90 (0.48-1.68) |
| No | 123 (35.55) | 223 (64.45) |  |
| CKD |  |  |  |
| Yes | 48 (30.38) | 110 (69.62) | 0.78 (0.58-1.04) |
| No | 82 (39.05) | 128 (60.95) |  |

#### 3.2 Foot care education

Seventy one percent of the patient declared that they had not received any information about foot care during consultation. the proportion of patients receiving foot education according to sociodemographic and clinical characteristics are reported in table 2. Information on foot care was more frequently provided in patient who were assessed by endocrinologist, those with longer diabetes duration and those who had history of DFU.

**Table 2.**
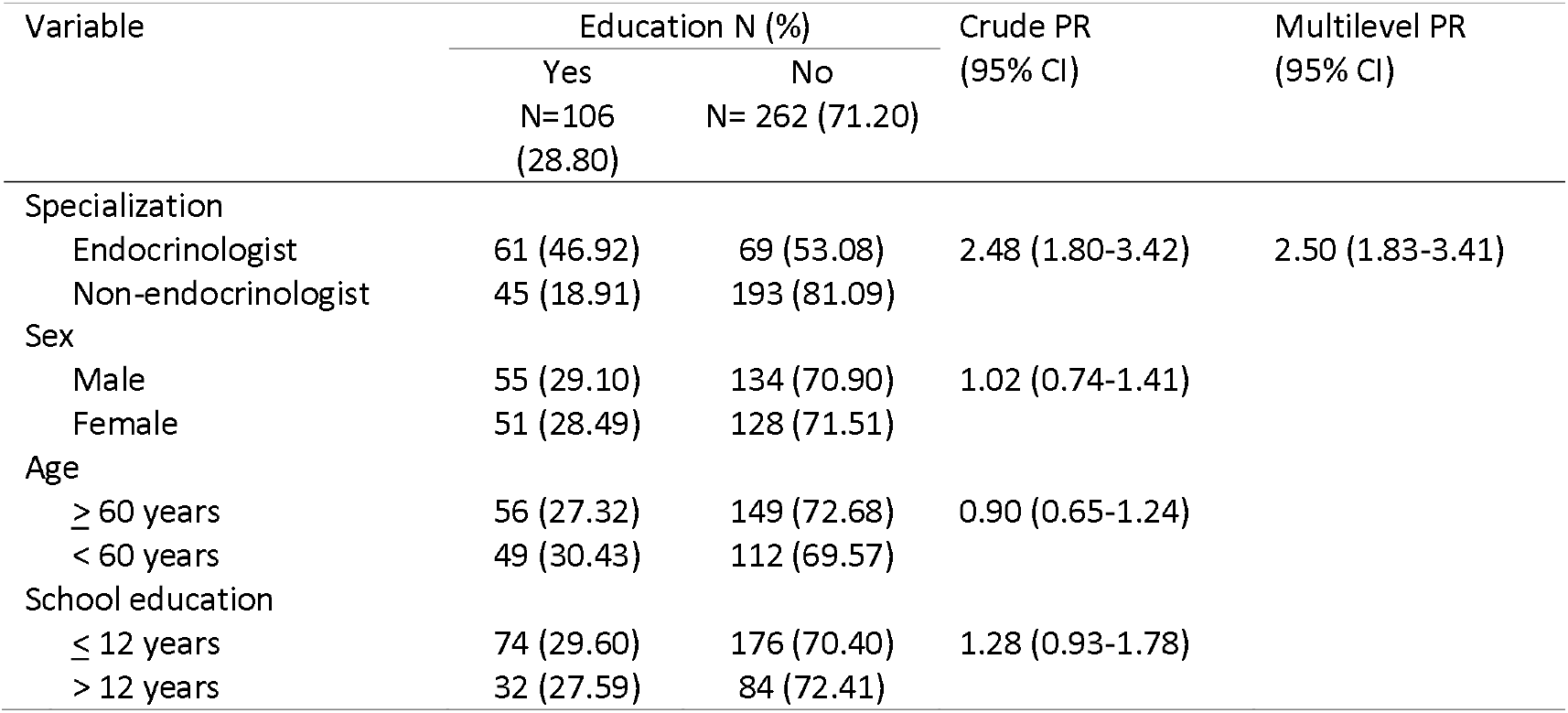
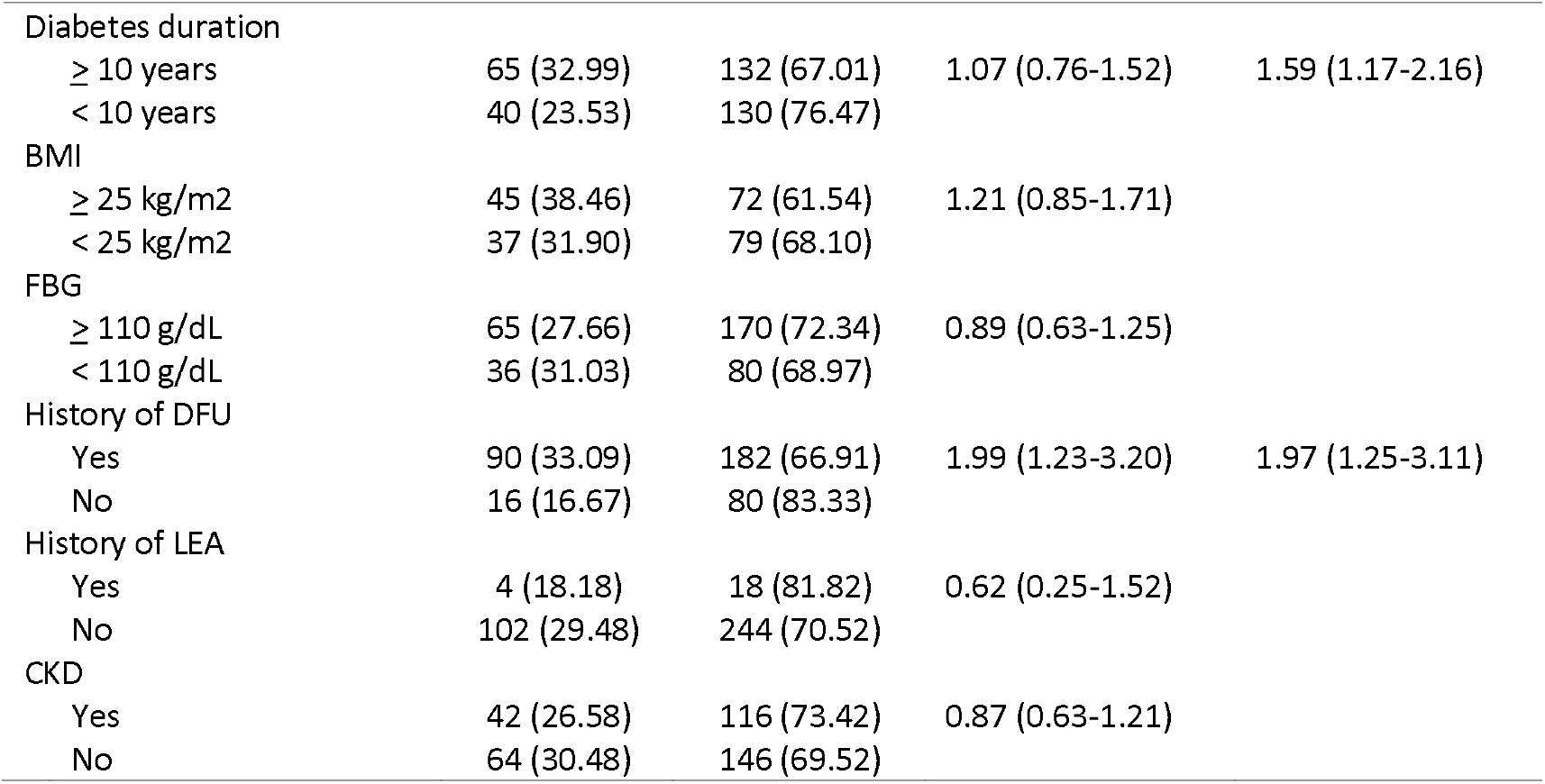
Foot education according to patient characteristics

### 3.3 Foot Examination

Overall, 169 patients (45.92%) declared that they prior internist had performed foot examination to them, while 199 (54.08%) declared that they had never received any foot examination according to their last consultation. The proportion of patient receiving foot examination according to sociodemographic, clinical characteristics and physician in charged are reported in table 3. Foot examination was more likely to performed in patient with longer diabetes duration and those who had history of LEA. Moreover, endocrinologist was more frequently perform foot examination compared to non-endocrinologist internist.

**Table 3.** Foot examination according to patient charateristics

| Variable | Foot Examination N (%) |  | Crude PR<br>(95% CI) | Multilevel PR (95%<br>CI) |
| --- | --- | --- | --- | --- |
|  | Yes<br>N=169<br>(45.92) | No<br>N=199<br>(54.08) |  |  |
| Specialty |  |  |  |  |
| Endocrinologist | 61 (46.92) | 69 (53.08) | 2.48 (1.80-3.42) | 2.19 (1.79-2.69) |
| Non-endocrinologist | 45 (18.91) | 193 (81.09) |  |  |
| Sex |  |  |  |  |
| Male | 94 (49.74) | 95 (50.26) | 1.19 (0.95-1.49) |  |
| Female | 75 (41.90) | 104 (58.10) |  |  |
| Age |  |  |  |  |
| ≥ 60 years | 99 (48.29) | 106 (51.71) | 1.13 (0.90-1.42) |  |
| < 60 years | 69 (42.86) | 92 (57.14) |  |  |
| School education |  |  |  |  |
| ≤ 12 years | 113 (45.20) | 137 (54.80) | 0.95 (0.75-1.21) |  |
| > 12 years | 55 (47.41) | 61 (52.59) |  |  |
| Diabetes duration |  |  |  |  |
| ≥ 10 years | 103 (52.28) | 94 (47.72) | 1.37 (1.08-1.73) | 1.45 (1.20-1.75) |
| < 10 years | 65 (38.24) | 105 (61.76) |  |  |
| BMI |  |  |  |  |
| ≥ 25 kg/m<sup>2</sup> | 62 (52.99) | 55 (47.01) | 1.23 (0.94-1.61) |  |
| < 25 kg/m<sup>2</sup> | 50 (43.10) | 66 (56.90) |  |  |
| FBG |  |  |  |  |
| ≥ 110 g/dL | 112 (47.66) | 123 (47.55) | 1.06 (0.83-1.36) |  |
| < 110 g/dL | 52 (44.83) | 64 (55.17) |  |  |
| History of DFU |  |  |  |  |
| Yes | 132 (48.53) | 140 (51.47) | 1.26 (0.95-1.67) |  |
| No | 37 (38.54) | 59 (61.46) |  |  |
| History of LEA |  |  |  |  |
| Yes | 11 (50.00) | 11 (50.00) | 1.10 (0.71-1.69) | 1.45 (1.29-1.62) |
| No | 158 (45.66) | 188 (54.34) |  |  |
| CKD |  |  |  |  |
| Yes | 75 (47.47) | 183 (52.53) | 1.06 (0.85-1.32) |  |
| No | 94 (44.76) | 116 (55.24) |  |  |

## 4. Discussion

To our knowledge, this is the first study evaluating internist’ practice towards foot care in individual with Type 2 Diabetes Mellitus in Indonesia especially in high risk patient. Our data show that internist attention to foot problems in high-risk diabetes patient is generally low, there are substantial proportion of patients who are not offered adequate foot care. In our findings about seventy percent of the high-risk patients did not received foot educations and most of them were treated by non-endocrinologist. The only patient characteristics associated with greater chance of giving foot education in our study was longer diabetes durations and history of DFU. People with longer diabetes duration (> 10 years) are at risk of developing foot ulcers, also a person with history of foot ulcer are at higher risk to develop recurrent foot ulcer.^9–11^ Therefore, author thought that attention to this particular person by internist are much higher. Same gloomy picture emerges when looking at internist practice, in our study only 45.92% of such patients had their feet examined during consultation, and most of the patients who were not examined were treated by non-endocrinologist. In line with our findings, a study from Italia showed that endocrinologist was performed significantly better in terms of foot examination compared to another physician.^12^ Also, those patients who received foot education and those who had their foot examined were at lower risk of not regularly checking their feet. Based on IWGDF recommendation, every person with diabetes should be examine annually for signs and symptoms of foot disease.^13^ Also, education should be given as it improves a patient’s foot self-care knowledge and self-protective behavior.^5^

Finally, some of the potential limitations of our study need to be discussed. First, patients were selected through consecutive sampling, this sampling methods could not be considered as representatives of the entire population. Second, in non-endocrinologist specialist, there are particularly among internist, who were more interested in diabetes care and in the training to become endocrinologist therefore they may not reflect the diabetes care delivered by non-endocrinologist.

## 5. Conclusion

It is important to increase among internist, the awareness for the prevention of diabetic foot. It is necessary to develop strategies to improve the care and awareness among health professional especially internist in treating patients with diabetes.

## Data Availability

The data that support the findings of this study are available from the corresponding author, IAK, upon reasonable request.

## Acknowledgment

The author wishes to acknowledge dr. Nissa, and dr. Kevin for their contribution to data acquisition.

## Notes

### Competing Interest Statement

The authors have declared no competing interest.

### Funding Statement

The authors received no financial support for the research, authorship, and/or publication of this article.

### Author Declarations

This study had been approved by ethical committee of Fatmawati General Hospital 021/KPP/XI/2016

